# Humoral immune responses to COVID-19 vaccination in people living with HIV receiving suppressive antiretroviral therapy

**DOI:** 10.1101/2021.10.03.21264320

**Authors:** Zabrina L. Brumme, Francis Mwimanzi, Hope R. Lapointe, Peter Cheung, Yurou Sang, Maggie C. Duncan, Fatima Yaseen, Olga Agafitei, Siobhan Ennis, Kurtis Ng, Simran Basra, Li Yi Lim, Rebecca Kalikawe, Sarah Speckmaier, Nadia Moran-Garcia, Landon Young, Hesham Ali, Bruce Ganase, Gisele Umviligihozo, F. Harrison Omondi, Kieran Atkinson, Hanwei Sudderuddin, Junine Toy, Paul Sereda, Laura Burns, Cecilia T. Costiniuk, Curtis Cooper, Aslam H. Anis, Victor Leung, Daniel Holmes, Mari L. DeMarco, Janet Simons, Malcolm Hedgcock, Marc G. Romney, Rolando Barrios, Silvia Guillemi, Chanson J. Brumme, Ralph Pantophlet, Julio S.G. Montaner, Masahiro Niikura, Marianne Harris, Mark Hull, Mark A. Brockman

**Author notes:** **Corresponding Author Contact Information:** Zabrina L. Brumme, Ph.D., Professor, Faculty of Health Sciences, Simon Fraser University, 8888 University Drive, Burnaby, BC, Canada, V5A 1S6. ZLB and MAB contributed equally.

## Abstract

Humoral responses to COVID-19 vaccines in people living with HIV (PLWH) remain incompletely understood. We measured circulating antibodies against the receptor-binding domain (RBD) of the SARS-CoV-2 spike protein, ACE2 displacement and live viral neutralization activities one month following the first and second COVID-19 vaccine doses in 100 adult PLWH and 152 controls. All PLWH were receiving suppressive antiretroviral therapy, with median CD4+ T-cell counts of 710 (IQR 525-935) cells/mm^3^. Nadir CD4+ T-cell counts ranged as low as <10 (median 280; IQR 120-490) cells/mm^3^. After adjustment for sociodemographic, health and vaccine-related variables, HIV infection was significantly associated with 0.2 log_10_ lower anti-RBD antibody concentrations (p=0.03) and ∼11% lower ACE2 displacement activity (p=0.02), but not lower viral neutralization (p=0.1) after one vaccine dose. Following two doses however, HIV was no longer significantly associated with the magnitude of any response measured. Rather, older age, a higher burden of chronic health conditions, and having received two ChAdOx1 doses (versus a heterologous or dual mRNA vaccine regimen) were independently associated with lower responses. After two vaccine doses, no significant correlation was observed between the most recent or nadir CD4+ T-cell counts and vaccine responses in PLWH. These results suggest that PLWH with well-controlled viral loads on antiretroviral therapy and CD4+ T-cell counts in a healthy range will generally not require a third COVID-19 vaccine dose as part of their initial immunization series, though other factors such as older age, co-morbidities, vaccine regimen type, and durability of vaccine responses will influence when this group may benefit from additional doses. Further studies of PLWH who are not receiving antiretroviral treatment and/or who have low CD4+ T-cell counts are needed.

## BACKGROUND

As people living with HIV (PLWH) may be at increased risk for severe COVID-19, possibly as a result of immunosuppression, higher rates of multi-morbidity and social determinants of health ^1-4^, COVID-19 vaccination is expected to benefit this group ^5^. Our understanding of immune responses to COVID-19 immunization in PLWH however remains limited, in part because relatively few PLWH were included in the clinical trials for the COVID-19 vaccines that have now been widely administered in Canada and Europe (∼196 for the BNT162b2 mRNA vaccine ^6,7^, 176 for the mRNA-1273 mRNA vaccine ^8^ and 54 and 103 PLWH respectively in the UK and South Africa for the ChAdOx1 viral vectored vaccine ^9^). Furthermore, immune response data from PLWH in these trials are currently only available for ChAdOx1 ^10,11^. “Real-world” COVID-19 vaccine immune response data from PLWH are also limited. While all three of these vaccines have shown effectiveness following their initial mass rollouts ^12-14^, and while clinical trial and observational data have shown robust vaccine-induced humoral immune responses in the general population ^15-17^, impaired responses have been reported in certain immunocompromised groups including solid organ transplant recipients ^18,19^, cancer patients ^20-22^, and individuals on immunosuppressive or immune-depleting therapies ^23-25^.

While antiretroviral therapy durably suppresses HIV to undetectable levels in plasma, restores CD4+ T-cell numbers, and can reverse HIV-induced immune dysfunction to a substantial extent ^26-29^, persistent immunopathology can nevertheless lead to blunting of immune responses to vaccination in PLWH ^30-32^. Though “real world” COVID-19 vaccine immunogenicity data in PLWH are emerging ^33-36^, these studies have featured limited numbers of PLWH and/or controls, and none have adjusted for chronic health conditions that may impair immune responses ^37^. Here, we characterize SARS-CoV-2-specific humoral immune responses after immunization with one and two doses of a COVID-19 vaccine in 100 PLWH and 152 control participants ranging from 22 to 88 years of age.

## RESULTS

### Cohort characteristics and COVID-19 vaccine rollout in British Columbia, Canada

Characteristics of the 100 PLWH and 152 controls are shown in **Table 1**. All PLWH were receiving antiretroviral therapy; the most recent plasma viral load, measured a median of 32 (Interquartile range [IQR] 7-54) days before enrolment, was <50 copies HIV RNA/mL for 95 PLWH, and between 71-162 copies/mL for the remaining five PLWH, though prior values were <50 copies/mL in all five of these cases. The most recent CD4+ T-cell count, measured a median of 44 (Interquartile range [IQR] 18-136) days before enrolment, was 710 (IQR 525-935; range 130-1800) cells/mm^3^. The estimated nadir CD4+ T-cell count, recorded a median of 8 (IQR 3.4-15) years before enrolment, was 280 (IQR 120-490; range <10-1010) cells/mm^3^.

**Table 1:**
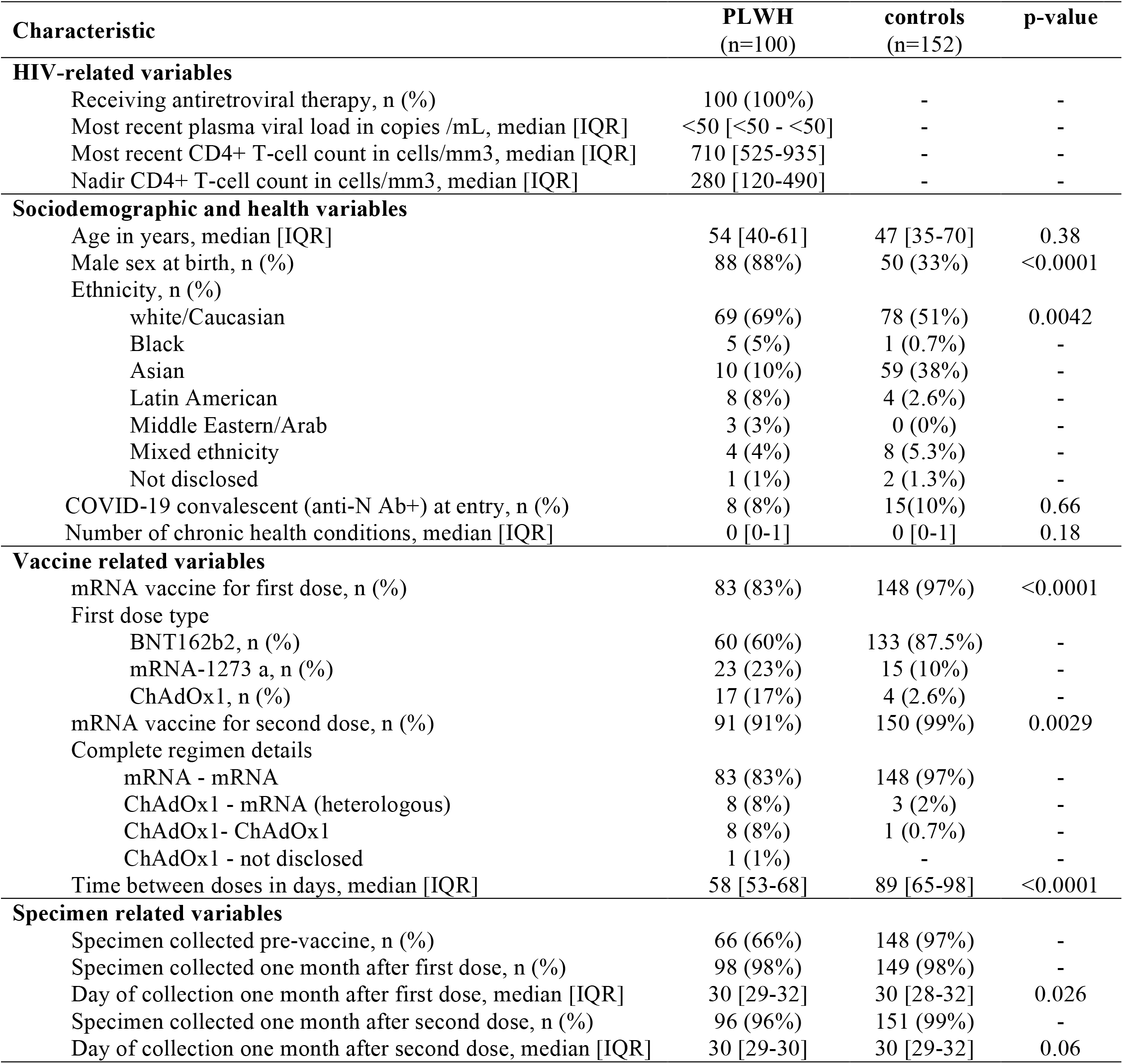
Participant characteristics.

| Characteristic | PLWH<br>(n=100) | controls<br>(n=152) | p-value |
| --- | --- | --- | --- |
| **HIV-related variables** |  |  |  |
| Receiving antiretroviral therapy, n (%) | 100 (100%) | - | - |
| Most recent plasma viral load in copies /mL, median [IQR] | <50 [<50 - <50] | - | - |
| Most recent CD4+ T-cell count in cells/mm3, median [IQR] | 710 [525-935] | - | - |
| Nadir CD4+ T-cell count in cells/mm3, median [IQR] | 280 [120-490] | - | - |
| **Sociodemographic and health variables** |  |  |  |
| Age in years, median [IQR] | 54 [40-61] | 47 [35-70] | 0.38 |
| Male sex at birth, n (%) | 88 (88%) | 50 (33%) | <0.0001 |
| Ethnicity, n (%) |  |  |  |
| white/Caucasian | 69 (69%) | 78 (51%) | 0.0042 |
| Black | 5 (5%) | 1 (0.7%) | - |
| Asian | 10 (10%) | 59 (38%) | - |
| Latin American | 8 (8%) | 4 (2.6%) | - |
| Middle Eastern/Arab | 3 (3%) | 0 (0%) | - |
| Mixed ethnicity | 4 (4%) | 8 (5.3%) | - |
| Not disclosed | 1 (1%) | 2 (1.3%) | - |
| COVID-19 convalescent (anti-N Ab+) at entry, n (%) | 8 (8%) | 15 (10%) | 0.66 |
| Number of chronic health conditions, median [IQR] | 0 [0-1] | 0 [0-1] | 0.18 |
| **Vaccine related variables** |  |  |  |
| mRNA vaccine for first dose, n (%) | 83 (83%) | 148 (97%) | <0.0001 |
| First dose type |  |  |  |
| BNT162b2, n (%) | 60 (60%) | 133 (87.5%) | - |
| mRNA-1273 a, n (%) | 23 (23%) | 15 (10%) | - |
| ChAdOx1, n (%) | 17 (17%) | 4 (2.6%) | - |
| mRNA vaccine for second dose, n (%) | 91 (91%) | 150 (99%) | 0.0029 |
| Complete regimen details |  |  |  |
| mRNA - mRNA | 83 (83%) | 148 (97%) | - |
| ChAdOx1 - mRNA (heterologous) | 8 (8%) | 3 (2%) | - |
| ChAdOx1 - ChAdOx1 | 8 (8%) | 1 (0.7%) | - |
| ChAdOx1 - not disclosed | 1 (1%) | - | - |
| Time between doses in days, median [IQR] | 58 [53-68] | 89 [65-98] | <0.0001 |
| **Specimen related variables** |  |  |  |
| Specimen collected pre-vaccine, n (%) | 66 (66%) | 148 (97%) | - |
| Specimen collected one month after first dose, n (%) | 98 (98%) | 149 (98%) | - |
| Day of collection one month after first dose, median [IQR] | 30 [29-32] | 30 [28-32] | 0.026 |
| Specimen collected one month after second dose, n (%) | 96 (96%) | 151 (99%) | - |
| Day of collection one month after second dose, median [IQR] | 30 [29-30] | 30 [29-32] | 0.06 |

PLWH and controls were similar in terms of age, but were different in terms of sex and ethnicity, with the PLWH group including more males and white ethnicity (**Table 1**). PLWH and controls had similar numbers of chronic health conditions (median 0; IQR 0-1; range 0-3 in both groups); the most common conditions were hypertension and asthma. At study entry, 8% of PLWH and 10% of controls were identified as COVID-19 convalescent based on the presence of anti-N antibodies. An additional one (1%) PLWH and two (1.5%) controls developed anti-N antibodies during follow-up consistent with SARS-CoV-2 infection after one vaccine dose. These participants were retained in the “COVID-19 naive at study entry” group, as excluding them did not affect results (not shown).

All participants received two COVID-19 vaccine doses between December 2020 and August 2021, with 97% of controls receiving an mRNA vaccine for their first dose compared to 83% of PLWH (**Table 1**). This is because health care workers, who represent 59% of controls, were eligible for vaccination before ChAdOx1 was approved in Canada, while members of the public, including PLWH, received the vaccine(s) recommended for their age group during the mass rollout. More PLWH received heterologous (ChAdOx1/mRNA) regimens compared to controls (8% and 2%, respectively). Heterologous regimens were administered in Canada after mRNA vaccines were universally recommended as second doses ^38^, after reports of rare thrombotic events associated with the ChAdOx1 vaccine emerged ^39^. The between-dose interval was also longer for the controls (median 89 days, versus 58 for PLWH). This is because the province of British Columbia (BC) extended the dose interval to 112 days beginning on March 1, 2021 due to limited vaccine supply ^40^, which meant that health care workers who were vaccinated around that time waited the longest for their second doses, while those vaccinated later waited a shorter time, as supplies increased. Samples were collected prior to vaccination where possible (66% of PLWH and 97% of controls), one month after the first vaccine dose (98% of both PLWH and controls) and one month after the second dose (96% of PLWH and 99% of controls).

### Anti-RBD binding antibody responses after first and second vaccine doses

Among participants naive to COVID-19 at study entry, all but three (one PLWH and two controls) developed anti-RBD antibodies after one vaccine dose, though overall concentrations in PLWH (median 1.50 [IQR 1.20-1.95] log_10_ U/mL) were on average ∼0.4 log_10_ lower than controls (median 1.94 [IQR 1.50-2.25] log_10_ U/mL); Mann-Whitney p=0.0001) (**Figure 1A and Supplemental Figure 1A**). In contrast, and consistent with prior studies demonstrating robust immune responses after one vaccine dose in previously infected individuals ^41,42^, anti-RBD antibody concentrations in COVID-19 convalescent participants (median 3.91 [IQR 3.21-4.26] log_10_ U/mL) were >2 log_10_ higher than in the COVID-19 naive PLWH or control participants (both p<0.0001) (convalescents were analyzed as a single group, since there was no statistically significant difference between PLWH and controls in this category; Mann-Whitney p=0.17).

**Figure 1:**
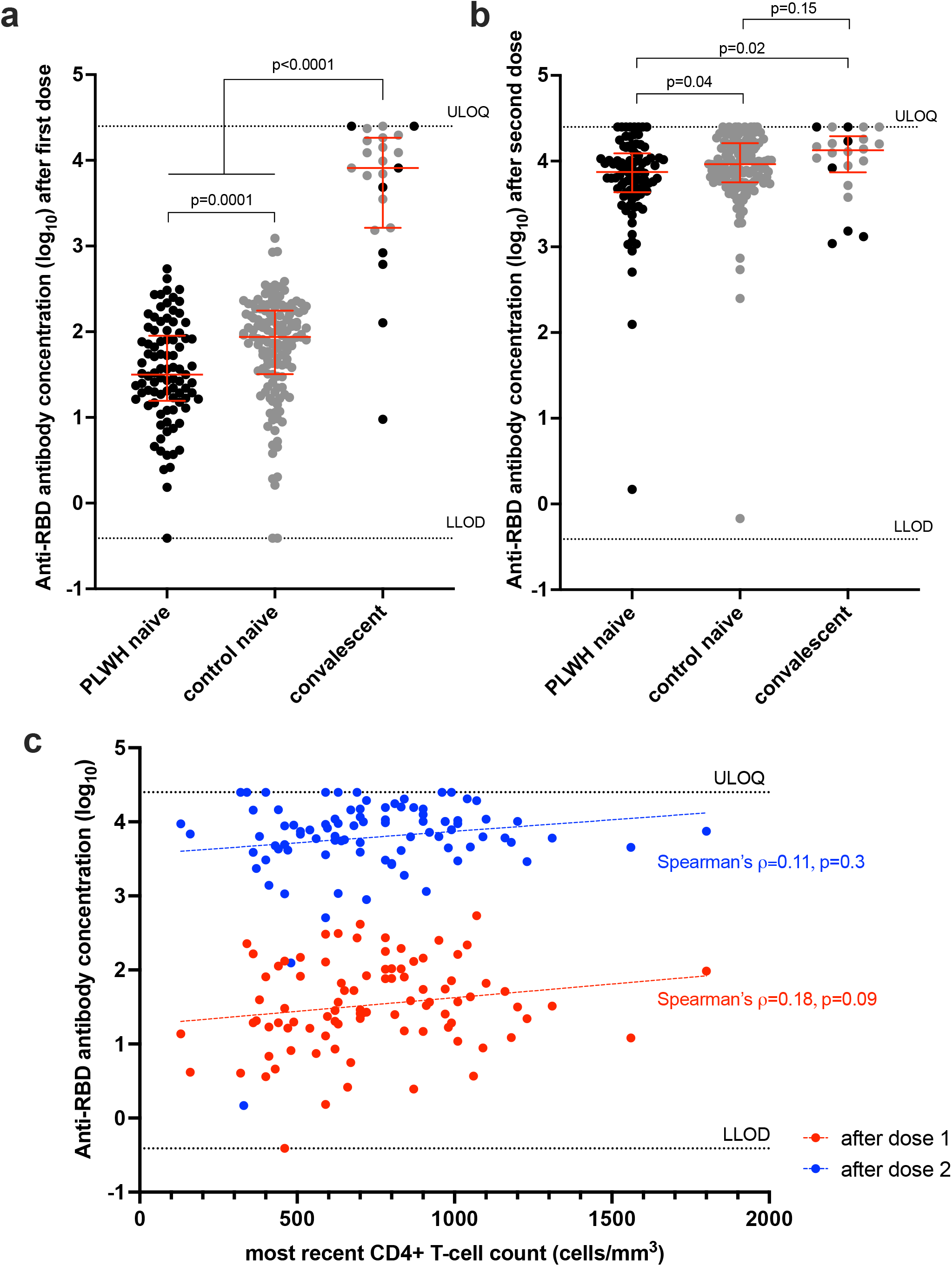
Binding antibody responses to spike RBD following one and two COVID-19 vaccine doses. *Panel A*: Binding antibody responses to the SARS-CoV-2 spike RBD in serum following one dose of a COVID-19 vaccine in PLWH (black circles) and controls (grey circles) who were COVID-19 naive at study entry. Convalescent participants, denoting those with anti-N antibodies at study entry, are colored as above. Red bars and whiskers represent the median and IQR. P-values were computed using the Mann-Whitney U-test and are uncorrected for multiple comparisons. LLOD: lower limit of detection. ULOQ: upper limit of quantification. *Panel B*: Binding antibody responses after two vaccine doses, colored as in A. *Panel C:* Correlation between most recent CD4+ T-cell count and binding antibody responses after one dose (red circles) and two doses (blue circles). Dotted lines are to help visualize the trend.

In multivariable analyses controlling for sociodemographic, health and vaccine-related variables, the strongest independent predictors of lower antibody responses after one dose were older age (every decade of age associated with an adjusted ∼0.1 log_10_ lower response; p=0.0002), and a higher number of chronic health conditions (every additional condition associated with an adjusted 0.14 log_10_ lower response; p=0.0058) (**Table 2**). HIV infection was also associated with an adjusted 0.2 log_10_ lower antibody response after one vaccine dose (p=0.031). Prior COVID-19 was associated with an adjusted 1.88 log_10_ higher response after one dose (p<0.0001).

**Table 2:**
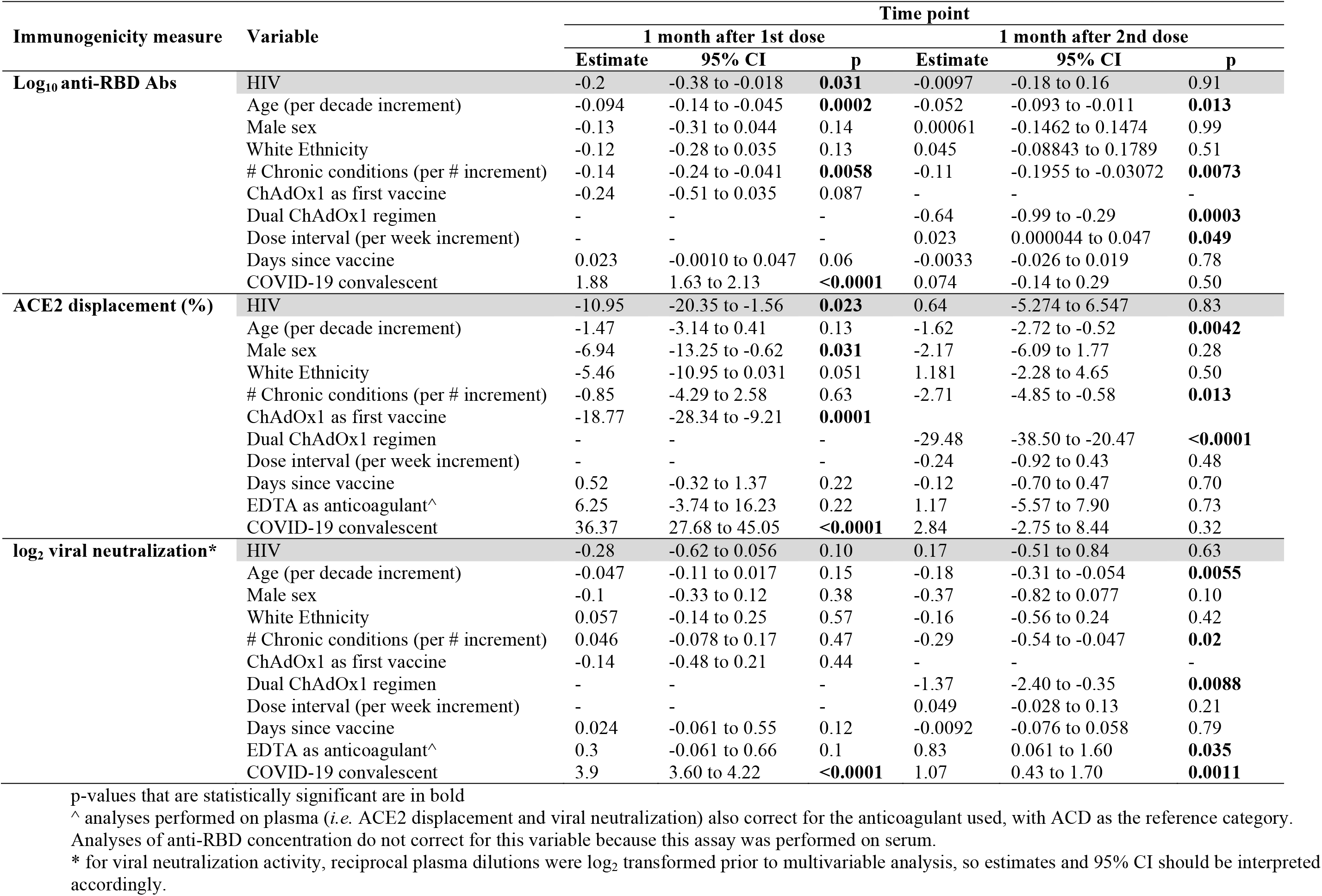
Multivariable analyses of the relationship between sociodemographic, health and vaccine-related variables on immunogenicity measures after first and second COVID-19 doses.

The second vaccine dose substantially boosted anti-RBD binding antibody concentrations in all but two participants: one PLWH with immunodeficiency due to a chronic blood disorder, and one >80 year old control participant with three chronic health conditions (**Figure 1B**). Overall, the second dose boosted anti-RBD levels in COVID-19 naïve individuals, both PLWH and controls, by a median of >2 log_10_, but in COVID-19 convalescent participants only marginally (median 0.14 log_10_), compared to that measured one month after the first dose (**Supplemental Figure 1B**). After two doses, antibody concentrations in COVID-19 naive PLWH (median 3.87 [IQR 3.64-4.09] log_10_ U/mL) were only ∼0.1 log_10_ lower than those in naive controls (median 3.96 [IQR 3.75-4.21] log_10_ U/mL; Mann-Whitney p=0.04), and only ∼0.2 log_10_ lower than in convalescent participants (median 4.13 [IQR 3.87-4.29] log_10_ U/mL; Mann-Whitney p=0.02) (**Figure 1B**).

In multivariable analyses, HIV infection was no longer associated with antibody concentrations after two vaccine doses (p=0.91, **Table 2**). Rather, older age, a greater number of chronic conditions and having received two ChAdOx1 doses were independently predictive of weaker responses, with every 10 years of older age, each additional chronic condition and having received dual ChAdOx1 doses associated with 0.052 log_10,_ 0.11 log_10_ and 0.64 log_10_ lower antibody concentrations, respectively (all p<0.02). A longer dose interval was also associated with marginally higher antibody concentrations (0.023 log_10_ per additional week, p=0.049). After two doses, there was no longer a significant association between prior COVID-19 infection and antibody response (p=0.50).

Among PLWH who were naive to COVID-19 at study entry, we observed a weak positive relationship between the most recent CD4+ T-cell count and antibody concentration after one dose that was not statistically significant (Spearman’s correlation ρ=0.18, p=0.09), but no significant relationship after the second dose (Spearman’s ρ=0.11, p=0.3; **Figure 1C**). Similarly we observed a weak positive relationship between *nadir* CD4+ T-cell count and antibody concentration after one dose that was not statistically significant (Spearman’s ρ=0.19, p=0.07), but no significant relationship after the second dose (Spearman’s ρ=0.05, p=0.6) (**Supplemental Figure 2A**).

**Figure 2:**
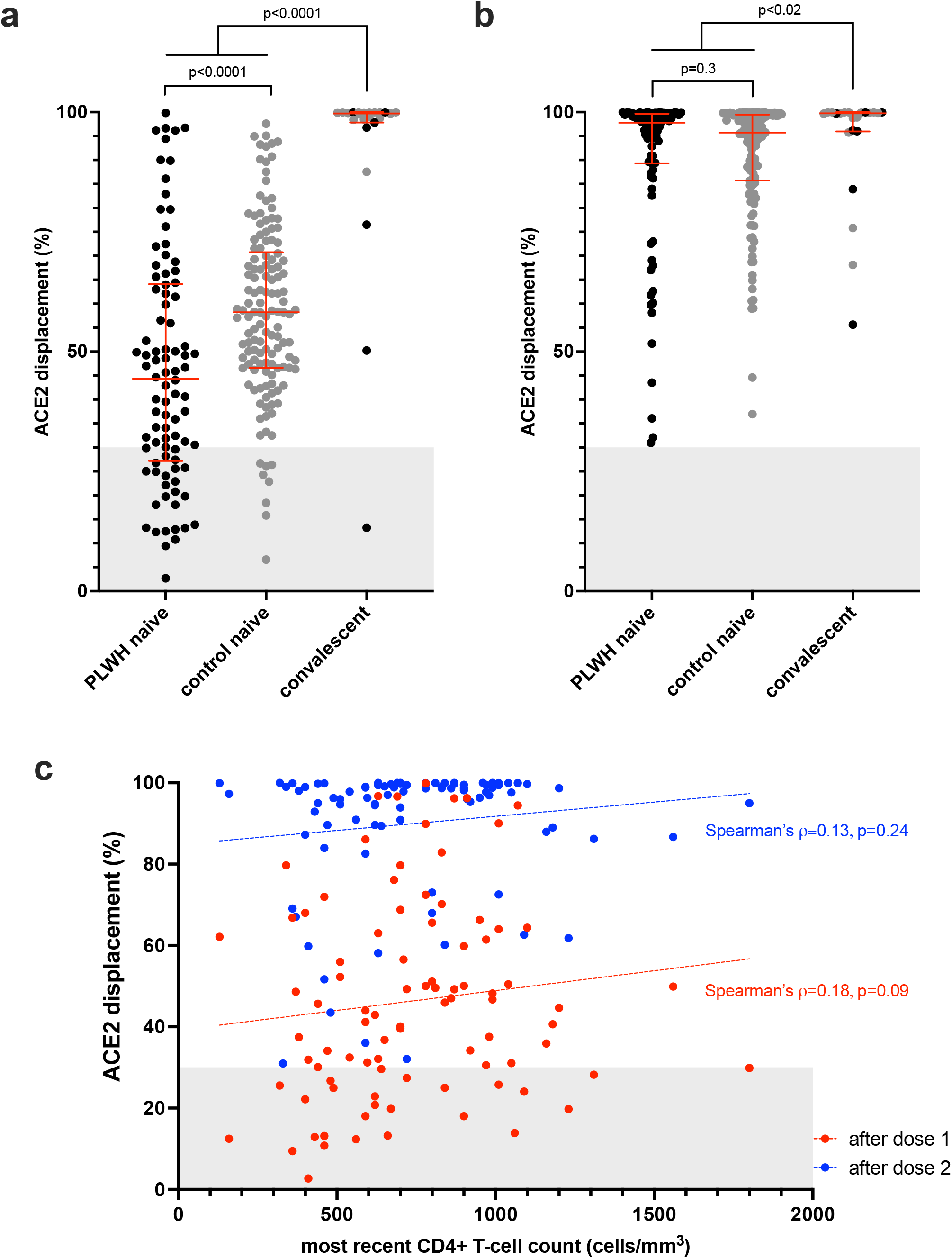
Ability of vaccine-induced antibodies to block ACE2-receptor binding following one and two COVID-19 vaccine doses. *Panel A*: ACE2 displacement activities of plasma antibodies following one dose of a COVID-19 vaccine in PLWH (black circles) and controls (grey circles) who were COVID-19 naive at study entry. Convalescent participants (those with anti-N antibodies at study entry) are colored as above. Red bars and whiskers represent median and IQR. Grey shaded area denotes the approximate range of values observed in pre-vaccine plasma from COVID-19 naive participants (see Supplemental Figure 1). P-values were computed using the Mann-Whitney U-test and are uncorrected for multiple comparisons. *Panel B*: ACE2 displacement activities after two vaccine doses, colored as in A. *Panel C:* Correlation between most recent CD4+ T-cell count and ACE2 displacement activities after one dose (red circles) and two doses (blue circles). Dotted lines are to help visualize the trend.

### ACE2 receptor displacement activities after first and second vaccine doses

We next assessed the ability of plasma to block the RBD-ACE2 interaction, which represents a higher throughput approach to estimate potential viral neutralization activity (also referred to as a surrogate viral neutralization test, sVNT ^43^). After one vaccine dose, PLWH and controls who were COVID-19 naive at study entry exhibited median 44% (IQR 27-64%) and 58% (IQR 47-71%) ACE2 displacement activities, respectively, indicating lower function among PLWH (Mann-Whitney p<0.0001) (**Figure 2A**). In contrast, convalescent participants exhibited a median 99.7% (IQR 97.8-99.9%) ACE2 displacement activity after one vaccine dose (Mann-Whitney p<0.0001 compared to both naive groups). In multivariable analyses, HIV infection remained significantly associated with an adjusted 11% lower ACE2 displacement activity after one vaccine dose (p=0.023), with male sex (adjusted ∼7% lower activity compared to female sex, p=0.031) and having received ChAdOx1 as the first dose (adjusted 18.8% lower activity compared to an mRNA vaccine as first dose, p=0.0001) remaining additional independent predictors of lower ACE2 displacement activity. Prior COVID-19 remained associated with an adjusted 36% higher ACE2 displacement activity following one vaccine dose (p<0.0001).

Following two vaccine doses, the median ACE2 displacement activity in COVID-19 naive PLWH and controls rose to >95% in both groups and there was no longer a statistically significant difference between them (median 97.8% [IQR 89.3-99.6%] in PLWH versus 95.7% [85.7%-99.5%] in controls, Mann-Whitney p=0.3) (**Figure 2B**). Furthermore, while the median ACE2 displacement activity in convalescent individuals (median 99.7% [IQR 96.0-100%]) remained statistically significantly higher than both naïve groups (both p<0.02), the magnitude of this difference was marginal. In fact, the second dose boosted ACE2 displacement activities in PLWH to an overall greater extent than in controls (**Supplemental Figure 1C, D**). Consistent with this, multivariable analyses identified older age, a larger number of chronic conditions, and dual ChAdOx1 vaccination – but not HIV – as being independently associated with lower ACE2 displacement function after two vaccine doses (adjusted 1.6% lower ACE2 displacement function for every decade of older age, 2.7% lower function for every additional health condition, and 29% lower function for dual ChAdOx1 vaccination; all p<0.02) (**Table 2**). Among PLWH who were naive to COVID-19 at study entry, we observed a weak positive correlation between recent CD4 + T-cell count and ACE2 displacement activity after one dose that was not statistically significant (Spearman’s ρ=0.18, p=0.09), and no association following two doses (Spearman’s ρ=0.13, p=0.24; **Figure 2C**). Similarly we observed a weak positive relationship between *nadir* CD4+ T-cell count and ACE2 displacement activity after one dose that was not statistically significant (Spearman’s ρ=0.2, p=0.06), but no significant relationship after the second dose (Spearman’s ρ=0.098, p=0.36) (**Supplemental Figure 2B)**.

### Viral neutralization activity after first and second vaccine doses

After one vaccine dose, plasma from most COVID-19 naive participants displayed weak or no ability to neutralize live SARS-CoV-2, with no significant differences between PLWH and controls (median/IQR undetectable in both groups; Mann-Whitney p=0.26) (**Figure 3A**). In contrast, neutralization activities in COVID-19 convalescent individuals were significantly higher, where reciprocal plasma dilutions needed to achieve neutralization were a median of 320 (IQR 80-320; p<0.0001 compared to both naive groups) (**Figure 3A**). Consistent with this, only COVID-19 convalescent status was significantly associated with higher neutralization activity in multivariable analyses after one dose (**Table 2**).

**Figure 3:**
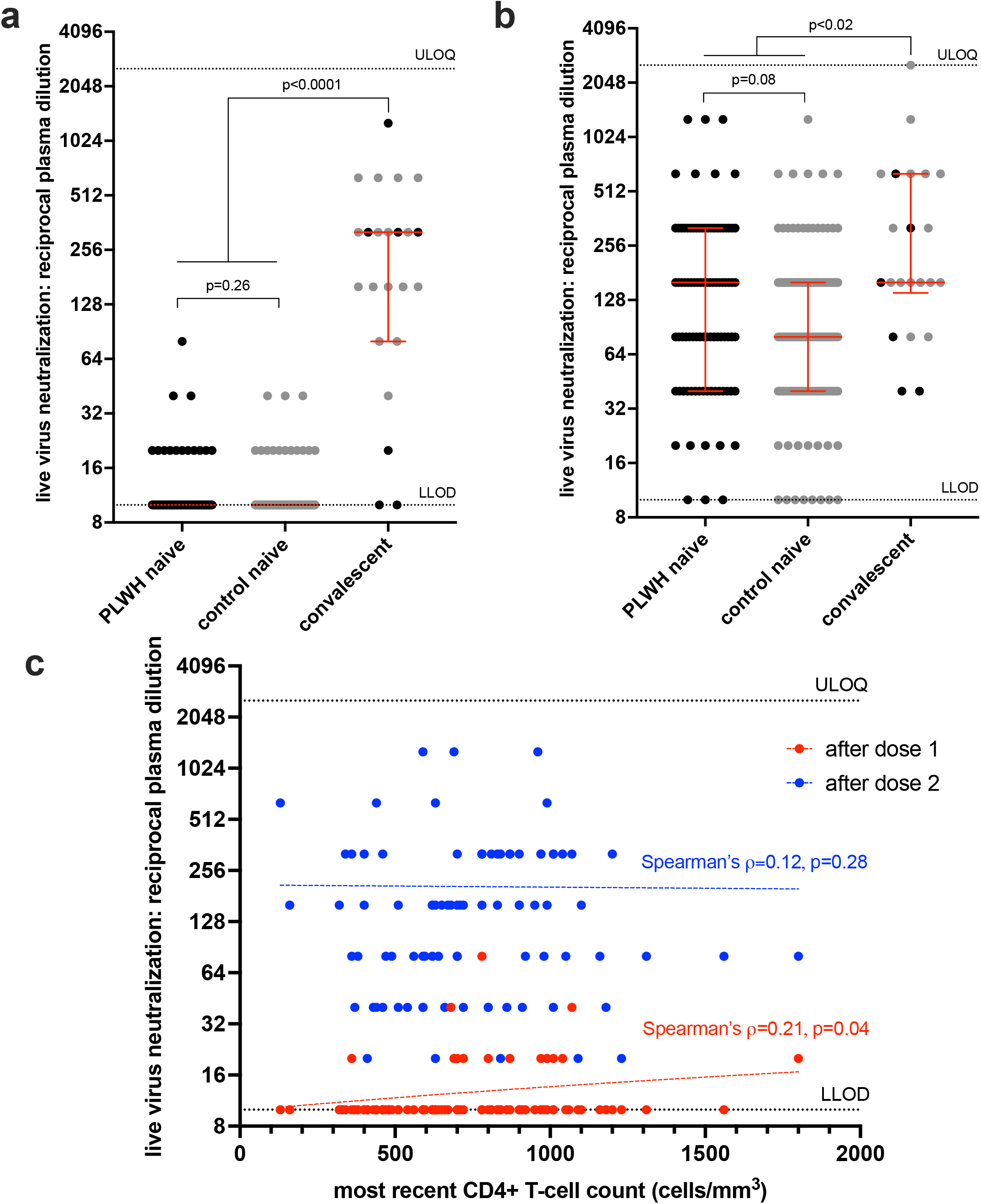
Ability of vaccine-induced antibodies to neutralize live SARS-CoV-2 following one and two COVID-19 vaccine doses. *Panel A*: Viral neutralization activities, defined as the lowest reciprocal plasma dilution at which neutralization was observed in all triplicate assay wells, following one dose of a COVID-19 vaccine in PLWH (black circles) and controls (grey circles) who were COVID-19 naive at study entry. Convalescent participants (those with anti-N antibodies at study entry, are colored as above. Red bars and whiskers represent median and IQR. P-values were computed using the Mann-Whitney U-test and are uncorrected for multiple comparisons. LLOD: assay lower limit of detection. ULOQ: assay upper limit of quantification. *Panel B*: Viral neutralization activities after two vaccine doses, colored as in A. *Panel C:* Correlation between most recent CD4+ T-cell count and viral neutralization activities after one dose (red circles) and two doses (blue circles). Dotted lines are to help visualize the trend.

Following two vaccine doses, viral neutralization activities in COVID-19 naive PLWH and controls increased an average of 8-fold in both groups (**Supplemental Figures 1E, D**), with naive PLWH achieving neutralization at median reciprocal plasma dilution of 160 (IQR 40-320) compared to a median of 80 (IQR 40-160) in controls (p=0.08) (**Figure 3B**). The viral neutralization activities of COVID-19 convalescent individuals (median reciprocal dilution of 160; IQR 140-640) remained marginally higher than COVID-19 naive individuals after two doses (p<0.02 for both comparisons). Consistent with the other humoral functions evaluated, multivariable analyses identified older age, a higher number of chronic conditions, and dual ChAdOx1 vaccination – but not HIV – as being independently associated with lower viral neutralization activity after two COVID-19 vaccine doses (p<0.02; **Table 2**).

Among PLWH who were naive to COVID-19 at study entry, we observed a weak positive correlation between recent CD4 + T-cell count and viral neutralization activity after one dose (Spearman’s ρ=0.21, p=0.04), but this association did not remain following two doses (Spearman’s ρ=0.12, p=0.28; **Figure 3C**). We observed no significant correlations between nadir CD4+ T-cell count and viral neutralization activity after either vaccine dose (**Supplemental Figure 2C)**.

### Humoral response against the SARS-CoV-2 delta variant

Given recent concerns that certain SARS-CoV-2 variants may be more transmissible or evade aspects of host immunity ^44,45^, we examined the ACE2 displacement activity in plasma against the widespread B.1.617.2 (Delta) variant. After one vaccine dose, plasma from all groups was impaired in its ability to block ACE2 receptor engagement by the Delta RBD compared to the original (Wuhan) RBD, where the magnitude of this impairment was a median of ∼8%, ∼19% and ∼1% for COVID-19 naive PLWH, naive controls and convalescents, respectively (Wilcoxon matched pairs signed rank test, all p≤0.0001) (**Figure 4A**). After two vaccine doses, these impairments remained, albeit at a much lower magnitude (a median of ∼2%, ∼8% and ∼1% for naive PLWH, naive controls and convalescents, respectively, all p<0.0001; **Figure 4B**). Given the strong correlations between ACE2 displacement and viral neutralization activities observed in our study (Spearman’s ρ≥0.58, p<0.0001; **Supplemental Figure 3**), these results suggest that vaccine-elicited humoral responses may be less able to prevent infection by the Delta variant, which is consistent with a recent report showing reduced ability of plasma from convalescent and vaccinated individuals to neutralize this strain ^46^.

**Figure 4:**
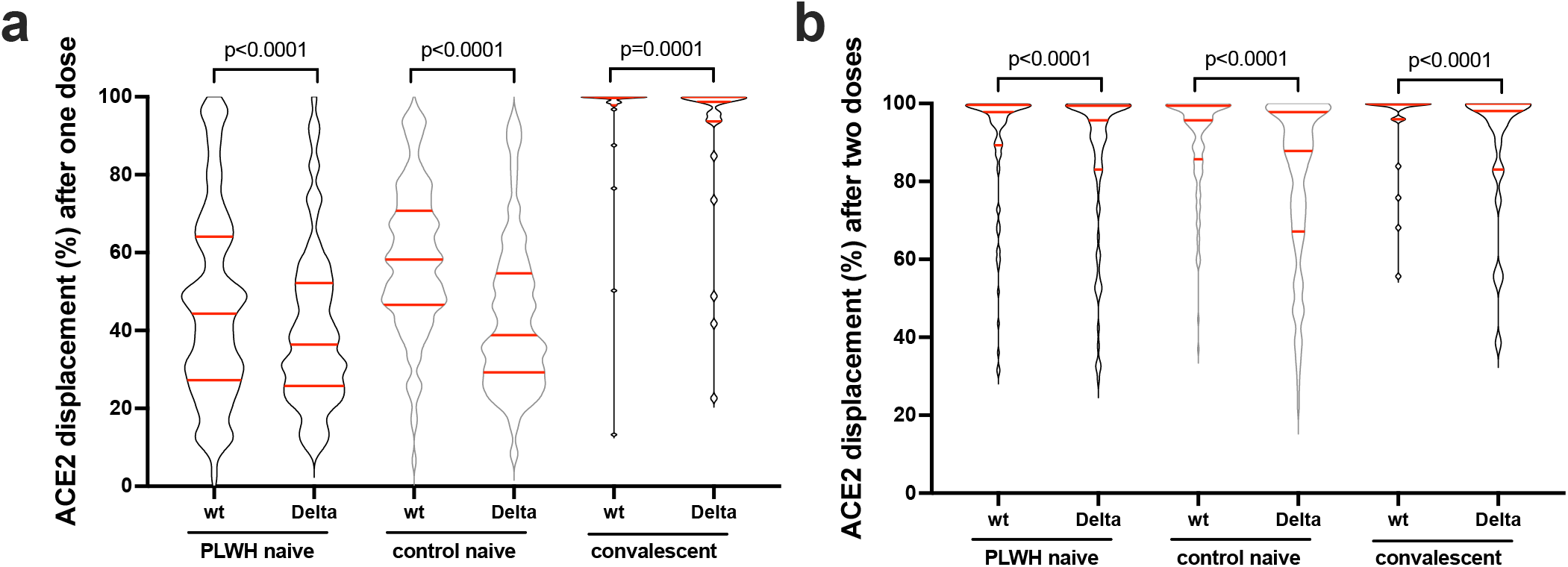
ACE2 displacement activities against the original and Delta SARS-CoV-2 variants after one and two doses of COVID-19 vaccine. *Panel A:* ACE2 displacement activities of plasma antibodies against the original wild-type (wt) and Delta variant Spike-RBD in naive PLWH, naive controls, and convalescent individuals after one vaccine dose. Data are shown as violin plots with horizontal red lines depicting the mean, 1st and 3rd quartiles. P-values were computed using the Wilcoxon matched-pairs signed rank test, and are uncorrected for multiple comparisons. *Panel B*: Same as panel A, but for responses after two vaccine doses.

## DISCUSSION

Our results add to a growing body of evidence that adult PLWH receiving stable antiretroviral therapy, who have suppressed plasma HIV loads and who have CD4+ T-cell counts in a healthy range, generally mount robust humoral immune responses to COVID-19 vaccines ^10,33-35^. Though HIV infection was associated with marginally (0.2 log_10_) lower overall anti-RBD antibody concentrations and ∼11% lower ACE2 displacement activities following a single vaccine dose after adjustment for sociodemographic, health and vaccine-related variables, we observed no effects of HIV infection on anti-RBD antibody concentrations, ACE2 displacement or viral neutralization activities after two vaccine doses. Rather, older age and a higher burden of chronic health conditions were independently associated with weaker humoral responses after two vaccine doses, consistent with previous reports ^37,47-50^. In addition, having received two ChAdOx1 doses, as opposed to a heterologous or autologous mRNA vaccine regimen, was associated with significantly lower “peak” humoral responses (measured one month following the second dose), which is also consistent with previous reports ^51,52^. Recent data however have indicated that, while humoral responses to the mRNA vaccines initially reach high levels but wane considerably thereafter, immune responses induced by a COVID-19 viral vectored vaccine induced lower median titers that remain more steady over time^53^. A longer interval between doses (where the maximum dose interval among study participants was 122 days) was also associated with marginally higher binding antibody concentrations, though not ACE2 displacement or viral neutralization activities, which is partially consistent with reports of improved antibody and T-cell responses using extended dosing intervals of the BNT162b2 mRNA vaccine ^54^. Indeed, Canada’s unique adoption of a very long (up to 112 days) interval between first and second COVID-19 vaccine doses yields insight into the magnitude of peak humoral responses following such an extended regimen. It is interesting that the anti-RBD antibody concentrations following two doses measured in the present study are generally higher than in studies of individuals who had shorter dose intervals that also employed the Roche Elecsys Anti-SARS-CoV-2 S assay, even though responses following dose 1 were similar ^55-58^. Comparing values across studies should be done with caution however, as the assay quantitative range will vary based on the maximum sample dilution performed.

Importantly, among PLWH in our study, all of whom were receiving suppressive antiretroviral treatment, we observed only a very weak positive correlation between the most recent CD4+ T-cell count and humoral responses after the first vaccine dose. Moreover, this association disappeared following the second vaccine dose. While CD4+ T-cell counts <250 cells/mm^3^ have been associated with lower antibody levels following one COVID-19 vaccine dose ^36^, we were unable to confirm this finding as only two PLWH in the present study had CD4+ T-cell counts in this range, and both of them mounted strong vaccine responses. Moreover, although we found weak positive correlations between *nadir* CD4+ T-cell counts (which were as low as <10 cells/mm^3^ in our cohort) and both anti-RBD antibody concentrations and ACE2 displacement activities following one dose, these associations no longer remained following the second dose. Furthermore, we observed no association between viral neutralization activity and nadir CD4 T+ cell count after either vaccine dose. This indicates that, for PLWH currently receiving suppressive antiretroviral therapy and whose CD4+ T-cell counts are currently in a healthy range, having had low CD4 T+ cell counts in the past will not necessarily compromise immune responses to COVID-19 vaccines presently.

We also observed that the ability of vaccine-induced plasma antibodies to disrupt the ACE2/RBD interaction was modestly yet significantly reduced against the RBD of the now widespread SARS-CoV-2 Delta variant compared to the original strain for all participant groups. Given the ability of SARS-CoV-2 variants to evade at least some aspects of vaccine-elicited immunity ^44^, this suggests that all individuals, regardless of HIV status, will remain more susceptible to infection by this variant, even after vaccination.

Our study has several limitations. Our results may not be generalizable to PLWH who are not receiving antiretroviral therapy and/or who have CD4+ T-cell counts <200 cells/mm^3^. Our study did not include children or adolescents living with HIV. As the precise immune correlates of protection for SARS-CoV-2 transmission and disease severity remain incompletely characterized ^59^, the implications of our results on individual-level protection from SARS-CoV-2 infection and COVID-19 remain uncertain. The relationship between vaccine-induced antibody concentrations in blood and at mucosal sites, which may be a better correlate of protection, is also incompletely understood, though a recent study identified anti-RBD IgG antibodies in saliva in 100% of participants following a two-dose COVID-19 vaccine series ^60^. We did not investigate vaccine-induced T-cell responses, though two recent studies have demonstrated comparable anti-Spike T-cell responses in PLWH compared to controls ^10,34^. Our study was not designed to investigate potential differences in immune responses between the two mRNA vaccines ^55,61,62^. The latest time-point we analyzed was one month after the second vaccine dose, so analyses of COVID-19 vaccine durability are also needed.

Taken together with existing data ^10,33-35^, our results indicate that adults with HIV receiving suppressive antiretroviral therapy, who have CD4+ T-cell counts in the healthy range mount broadly comparable “peak” humoral immune responses to two COVID-19 vaccine doses compared to individuals without HIV. Furthermore, we found no evidence that a low nadir CD4+ T-cell count negatively influenced the response to COVID-19 vaccination in this group. Rather, our results identified older age, additional chronic health conditions, and having received a dual ChAdOx1 regimen (as opposed to a heterologous or dual mRNA vaccine regimen) – but not HIV – as negative modulators of humoral responses following two-dose COVID-19 vaccination.

These results suggest that PLWH whose viral loads are well-controlled on antiretroviral therapy and whose CD4+ T-cell counts are in a healthy range will generally not require a third COVID-19 vaccine dose as part of their initial immunization series, though other factors such as older age, co-morbidities, type of initial vaccine regimen and durability of vaccine responses will influence when this group may benefit from additional doses. Further studies of PLWH who are not receiving antiretroviral treatment and/or who have low CD4+ T-cell counts are needed.

## METHODS

### Participants and sampling

A total of 100 adult PLWH were recruited through three HIV care clinics in Vancouver, British Columbia (BC), Canada and through community outreach. A total of 152 control participants without HIV included 24 adults <65 years of age recruited for the present study, along with 39 community-dwelling adults >65 years of age and 89 health care workers who were recruited for a parallel study of COVID-19 vaccine immune responses across the adult age spectrum ^63^. HIV-negative status of control participants was determined by self-report. Serum and plasma were collected prior to COVID-19 vaccination (where possible), one month after the first COVID-19 vaccine dose, and at one month after the second dose. Plasma was collected in either ethylenediaminetetraacetic acid (EDTA) or anticoagulant citrate dextrose (ACD). Specimens were processed on the day of collection and frozen until analysis. COVID-19 convalescent individuals were identified at study entry by the presence of serum antibodies against the SARS-CoV-2 nucleoprotein (N).

### Ethics approval

This study was approved by the University of British Columbia/Providence Health Care and Simon Fraser University Research Ethics Boards. All participants provided written informed consent.

### Data sources

Sociodemographic data, chronic health conditions and COVID-19 vaccination information were collected by self-report and confirmed through medical records where available. We assigned a score of 1 for each of the following 11 chronic health conditions: hypertension, diabetes, asthma, obesity (defined as having a body mass index ≥30), chronic diseases of lung, liver, kidney, heart or blood, cancer, and immunosuppression due to chronic conditions or medication, to generate a total score ranging from 0-11. Clinical information for PLWH was recovered from the BC Centre for Excellence in HIV/AIDS Drug Treatment Program Database, which houses clinical records for all PLWH receiving care in BC. For PLWH, having a recent CD4+ T-cell count <200 cells/mm^3^ was classified as “immunosuppression” in the chronic health conditions score.

### Binding antibody assays

We measured total binding antibodies against SARS-CoV-2 N and spike receptor binding domain (RBD) in serum using the Roche Elecsys Anti-SARS-CoV-2 and Elecsys Anti-SARS-CoV-2 S assays, respectively. Post-infection, both anti-N and anti-RBD assays should be positive, whereas post-mRNA vaccination only the anti-RBD should be positive, enabling identification of convalescent samples. Both assays are electro-chemiluminescence sandwich immunoassays, and report results in arbitrary units/mL (U/mL), calibrated against an external standard. For the S assay, the manufacturer indicates that AU values can be considered equivalent to international binding antibody units (BAU) as defined by the World Health Organization and the measurement range is from 0.4 - 25,000 U/mL ^64^.

### ACE2 displacement assay

We assessed the ability of plasma antibodies to block the interaction between RBD and the ACE2 receptor using the V-plex SARS-CoV-2 Panel 11 (ACE2) kit on a MESO QuickPlex SQ120 instrument (Meso Scale Discovery) at the manufacturer’s recommended 1:20 dilution. ACE2 displacement was calculated as 100 - [Arbitrary Units (AU) of ACE2 binding in the presence of plasma / AU of ACE2 binding in the absence of plasma] and reported as a percentage.

### Live virus neutralization assay

Neutralizing activity in plasma was examined using a live SARS-CoV-2 infectivity assay in a Containment Level 3 facility. Assays were performed using isolate USA-WA1/2020 (BEI Resources) and VeroE6-TMPRSS2 (JCRB-1819) target cells. Viral stock was adjusted to 50 TCID_50_/200 µl in DMEM in the presence of serial 2-fold dilutions of plasma (from 1/20 to 1/2560), incubated at 4°C for 1 hour and then added to target cells in 96-well plates in triplicate. Cultures were maintained at 37°C with 5% CO_2_ and the appearance of viral cytopathic effect (CPE) was recorded 3 days post-infection. Neutralizing activity is reported as the reciprocal plasma dilution necessary to prevent CPE in all three replicate wells. Samples exhibiting only partial or no neutralization at 1/20 were coded as having a reciprocal dilution of “10”, defined as below the limit of detection in this assay.

### Statistical analysis

Comparisons of binary variables between groups were performed using Fisher’s exact test. Comparisons of continuous variables between groups were performed using the Mann-Whitney U-test (for unpaired data) or Wilcoxon test (for paired data). Correlations between continuous variables were performed using Spearman’s correlation. Multiple linear regression was employed to investigate the relationship between sociodemographic, health and vaccine-related variables and humoral outcomes. Analyses performed following one dose included age (per decade increment), sex at birth (female as reference group), ethnicity (non-white as reference group), number of chronic health conditions (per number increment), sampling date following vaccine dose (per day increment), and type of vaccine received (mRNA vaccine as reference group). Analyses performed following two doses additionally included the interval between doses (per week increment) and having received two ChAdOx1 doses (versus having received a heterologous or dual mRNA vaccine regimen). For assays that tested plasma (ACE2 displacement and viral neutralization), models also corrected for the anticoagulant used (ACD as the reference group). All tests were two-tailed, with p=0.05 considered statistically significant. Analyses were conducted using Prism v9.2.0 (GraphPad).

## Data Availability

Data are available upon reasonable request to the corresponding author. As per funder requirements, all data will also be deposited into a national database at the conclusion of the study.

## ACKNOWLEDGEMENTS

We thank the leadership and staff of Providence Health Care for their support of this study. We thank the phlebotomists and laboratory staff at St. Paul’s Hospital, the BC Centre for Excellence in HIV/AIDS and Simon Fraser University for assistance. Above all, we thank the participants, without whom this study would not have been possible.

This work was supported by funding from Genome BC, the Michael Smith Foundation for Health Research, and the BCCDC Foundation for Public Health through a rapid SARS-CoV-2 vaccine research initiative in BC award (VAC-009 to ZLB, MAB). It was also supported by the Public Health Agency of Canada (PHAC) through two COVID-19 Immunology Task Force (CITF) COVID-19 Awards (to ZLB, MGR, MAB and to CTC, CC, AHA), the Canada Foundation for Innovation through Exceptional Opportunities Fund – COVID-19 awards (to CJB, MAB, MN, MLD, RP, ZLB), a British Columbia Ministry of Health–Providence Health Care Research Institute COVID-19 Research Priorities Grant (to CJB), the CIHR Canadian HIV Trials Network (CTN) (to AHA) and the National Institute of Allergy and Infectious Diseases of the National Institutes of Health (R01AI134229 to RP). MLD and ZLB hold Scholar Awards from the Michael Smith Foundation for Health Research. LYL was supported by an SFU Undergraduate Research Award. GU and FHO are supported by Ph.D. fellowships from the Sub-Saharan African Network for TB/HIV Research Excellence (SANTHE), a DELTAS Africa Initiative [grant # DEL-15-006]. The DELTAS Africa Initiative is an independent funding scheme of the African Academy of Sciences (AAS)’s Alliance for Accelerating Excellence in Science in Africa (AESA) and supported by the New Partnership for Africa’s Development Planning and Coordinating Agency (NEPAD Agency) with funding from the Wellcome Trust [grant # 107752/Z/15/Z] and the UK government. HS is supported by a CGS-M award from the Canadian Institutes of Health Research (CIHR). The views expressed in this publication are those of the authors and not necessarily those of PHAC, CITF, AAS, NEPAD Agency, Wellcome Trust, the Canadian or UK governments or other funders.

## AUTHOR CONTRIBUTIONS

Z.L.B and M.A.B. conceived and oversaw the study, drafted the manuscript and acquired funding. H.L. coordinated study operations, and collected, managed, curated and analyzed data. F.M., P.C., Y.S., M.C.D., F.Y, O.A, S.E., K.N., S.B, L.Y.L, R.K., S.S., N.M.-G, L.Y. B.G., G.U., F.H.O., K.A., H.S., and L.B. collected and/or analyzed data. J.T. and P.S. curated data. D.H., M.L.D., J.S., and M.G.R oversaw the collection of the Roche Elecsys data and provided technical guidance and expertise. H.A., V.L., D.H., M.L.D., J.S., M.H., M.G.R, R.B., S.G., J.S.G., M.H. and M.H provided clinical expertise, guidance and input. C.J.B provided input on statistical analysis and analyzed data. M.N., and R.P. designed assays, conducted/oversaw data collection and provided guidance and input. C.T.C., C.C. and A.H.A. provided guidance and input and acquired additional funding. All authors discussed the results and implications and commented on the manuscript at all stages.

