## Supplemental Figures for "Humoral immune responses to COVID-19 vaccination in people living with HIV receiving suppressive antiretroviral therapy"

Supplemental Figure 1

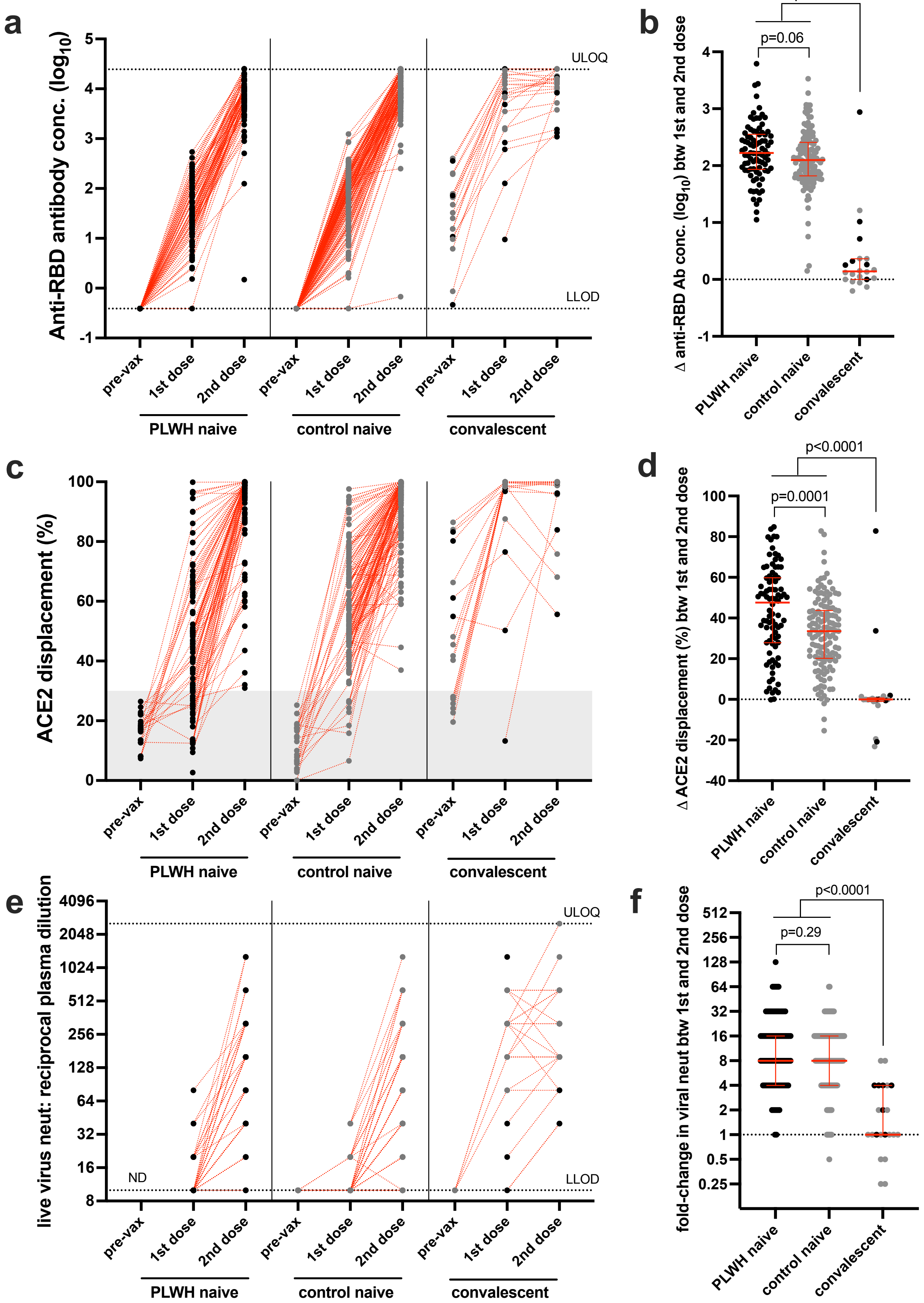

**Supplemental Figure 1 (previous page): Humoral immune response measurements pre- and post-vaccination** *Panel A:* Binding antibody responses in COVID-19 naive PLWH (black circles), COVID-19 naive controls (grey circles) and COVID-19 convalescent individuals (colored as above) pre- and post-vaccination. Red dotted lines connect participants' longitudinal measurements. LLOD: lower limit of detection. ULOQ: upper limit of quantification. *Panel B*: Difference in $\log_{10}$ binding antibody concentration between the first and second vaccine doses, with groups colored as above. P-values are computed using the Mann-Whitney U test, and are uncorrected for multiple comparisons. *Panel C*: Same as A, except showing ACE2 displacement activity. Pre-vaccine measurements were only performed on a subset of COVID-19 naive participants to estimate assay background, shown as grey shading. *Panel D:* Same as B, except showing ACE2 displacement activity. *Panel E*: Same as A, except showing viral neutralization activity. Pre-vaccine measurements were only performed on a subset of COVID-19 naive controls and convalescent individuals, none of whom had detectable neutralization activity at this time. ND=not determined. *Panel F:* Similar to B, except for viral neutralization activity, which is depicted as the fold-change in activity between 1st and 2nd doses.

**Supplemental Figure 2: Correlation between nadir CD4+ T-cell count and COVID-19 vaccine responses**

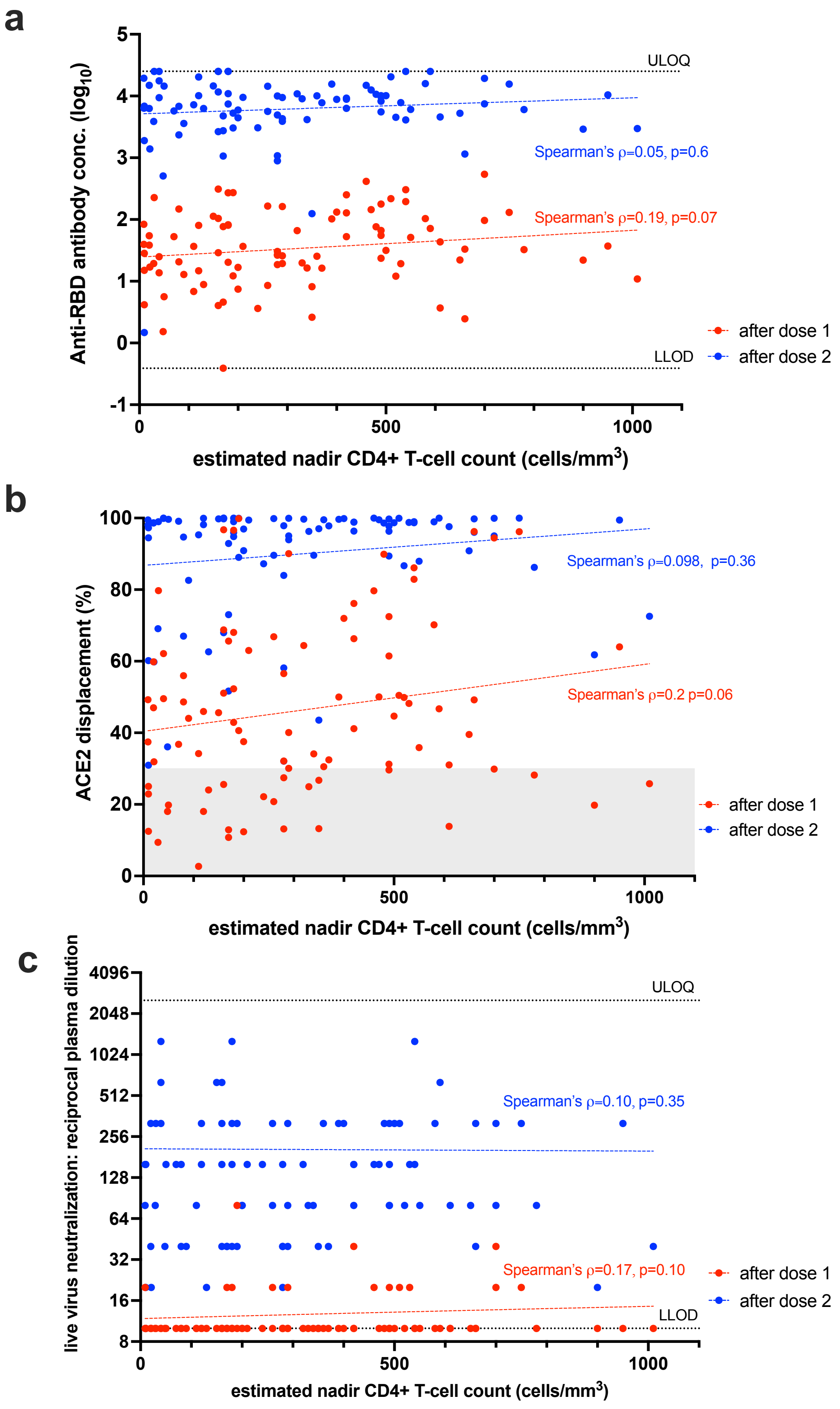

**Supplemental Figure 2 (previous page): Correlation between nadir CD4+ T-cell count and COVID-19 vaccine responses.** *Panel A:* Correlation between nadir CD4+ T-cell count and binding antibody responses after one dose (red circles) and two doses (blue circles). Dotted lines are to help visualize the trend. LLOD: lower limit of detection. ULOQ: upper limit of quantification. *Panel B*: same as A, but showing ACE2 displacement activity. Grey shading denotes approximate assay background levels, determined by testing pre-vaccine samples from COVID-19 naive individuals. *Panel C*: same as A, but showing viral neutralization activity.

**Supplemental Figure 3: Correlations between antibody responses/activities after one and two doses of COVID-19 vaccine**

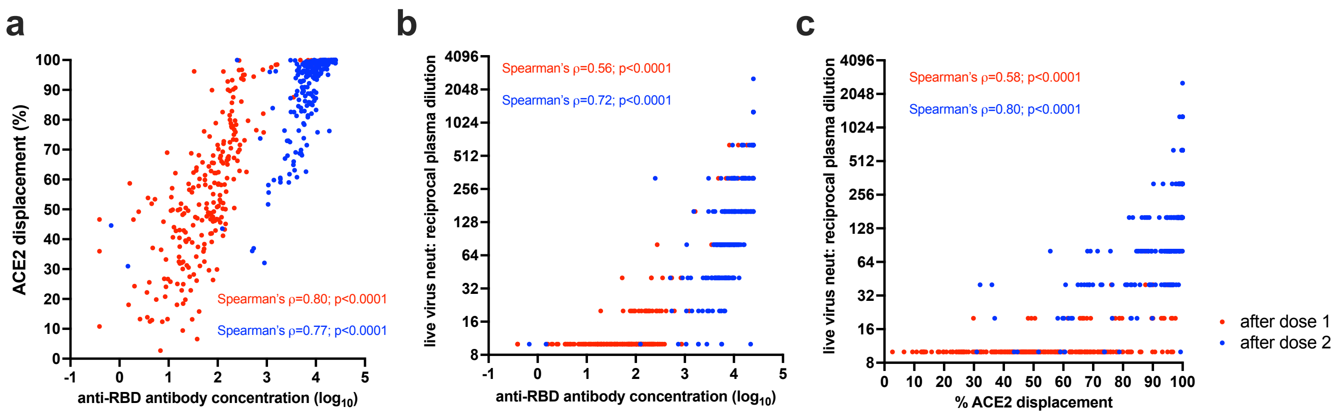
